# The PAX1 locus at 20p11 is a modifier for bilateral cleft lip only

**DOI:** 10.1101/2020.11.30.20241141

**Authors:** Sarah W. Curtis, Daniel Chang, Myoung Keun Lee, John R. Shaffer, Karlijne Indencleef, Michael P. Epstein, David J. Cutler, Jeffrey C. Murray, Eleanor Feingold, Terri H. Beaty, Peter Claes, Seth M. Weinberg, Mary L. Marazita, Jenna C. Carlson, Elizabeth J. Leslie

## Abstract

Nonsyndromic orofacial clefts (OFCs) are the most common craniofacial birth defect in humans and, like many complex traits, OFCs are phenotypically and etiologically heterogenous. The phenotypic heterogeneity of OFCs extends beyond the structures affected by the cleft (e.g., cleft lip (CL) and cleft lip and palate (CLP) to other features, such as the severity of the cleft. Here, we focus on bilateral and unilateral clefts as one dimension of OFC severity. Unilateral clefts are more frequent than bilateral clefts for both CL and CLP, but the genetic architecture of these subtypes is not well understood, and it is not known if genetic variants predispose for the formation of one subtype over another. Therefore, we tested for subtype-specific genetic associations in 44 bilateral CL (BCL) cases, 434 unilateral CL (UCL) cases, 530 bilateral CLP cases (BCLP), 1123 unilateral CLP (UCLP) cases, and unrelated controls (N = 1626), using the mixed-model approach implemented in GENESIS. While no novel loci were found in subtype-specific analyses comparing cases to controls, the genetic architecture of UCL was distinct compared to BCL, with 43.8% of suggestive loci (p < 1.0×10^−5^) having non-overlapping confidence intervals between the two subtypes. To further understand the genetic risk factors for severity differences, we then performed a genome-wide scan for modifiers using a similar mixed-model approach and found one genome-wide significant modifier locus on 20p11 (p = 7.53×10^−9^), 300kb downstream of *PAX1*, associated with higher odds of BCL compared to UCL, which also replicated in an independent cohort (p = 0.0018) and showed no effect in BCLP (p>0.05). We further found that SNPs at this locus were associated with normal human nasal shape. Taken together, these results suggest bilateral and unilateral clefts may have differences in their genetic architecture, especially between CL and CLP. Moreover, our results suggest BCL, the rarest form of OFC, may be genetically distinct from the other OFC subtypes. This expands our understanding of genetic modifiers for subtypes of OFCs and further elucidates the genetic mechanisms behind the phenotypic heterogeneity in OFCs.

## Introduction

Orofacial clefts (OFCs), including cleft lip (CL), cleft lip and palate (CLP) and cleft palate (CP), are common, complex birth defects. Affecting 1 in 700 births worldwide, they are caused when one or more of the developmental programs during the first seven weeks of pregnancy that determine the form the face do not occur properly (1). While some OFCs present in conjunction with other congenital abnormalities, a majority of OFCs are classified as isolated, nonsyndromic (nsOFC), which are caused by a complex combination of genetic and environmental factors and have been the focus of numerous genome-wide association studies (2-13).

OFCs have striking phenotypic heterogeneity. OFCs are typically categorized into three subtypes: cleft lip only (CL), cleft lip and palate (CLP), and cleft palate only (CP) where CL includes clefts confined to the primary palate, CLP includes clefts that extend into the secondary palate, and CP affects the secondary palate only. CL and CLP are often combined into a more general category of cleft lip with or without cleft palate (CL/P) based on the shared defect of the primary palate. OFCs affecting the primary palate can also be further subdivided based on morphological details to capture severity, including the laterality (unilateral or bilateral), the side of unilateral clefts (left or right), or the completeness of the cleft.

Population-based studies estimating recurrence risks have focused on different classifications of OFCs, and the resulting estimates can inform genetic models and the design of association studies. For example, among CLP cases, there is no difference in the risk of either CL or CLP among their first-degree relatives; suggesting a shared genetic etiology (14, 15) contributing to the rationale of studying CL/P in genetic association studies and many of the known risk loci show similar effects between CL and CLP (2-4, 16) (17).

Less is known about severity in CL and CLP or if there is a separate genetic component to cleft lip severity. Recurrence risk estimates based on severity are limited by sample size and have yielded mixed results. Semi-quantitative measures of completeness showed no effect of severity on estimated recurrence risks (15). However, the recurrence risk for bilateral clefts is higher than for unilateral clefts, indicating this more severe cleft type tends to recur more often in family members (14). Previous studies examining genetic factors associated with bilateral vs. unilateral clefts have been limited to targeted sequencing of a few selected candidate loci (17), although this work has suggested the presence of a genetic contribution to the different subtypes of cleft lip. Therefore, we set out to perform a genome-wide association study (GWAS) to determine if there are additional genetic variants that modify cleft severity by focusing on bilateral and unilateral clefting in CL and CLP cases.

## Methods

### Sample collection and SNP quality control

This study used samples from the Pittsburgh Orofacial Cleft (POFC) Study. The details of the sample collection and genotype quality control (QC) have been described previously (2, 18-20). Briefly, these samples came from 18 sites in 13 countries, including in the continental United States, Guatemala, Argentina, Colombia, Puerto Rico, China, Philippines, Denmark, Turkey, and Spain. All sites had Institutional Review Board (IRB) approval, both locally and at the University of Pittsburgh or University of Iowa, for genomic studies and data sharing. The original study recruited individuals with OFCs, their unaffected relatives, and unrelated controls (individuals with no known family history of OFCs or other craniofacial anomalies; N = 1626). For the current study, affected individuals were classified as either having a bilateral cleft lip (BCL; N = 44), a bilateral cleft lip and palate (BCLP; N = 530), a unilateral cleft lip (UCL; N = 434), or a unilateral cleft lip and palate (UCLP; N = 1123). Although this sample was not recruited with a population-based approach, the relative frequencies of these cleft types in the POFC study are consistent with epidemiological reports of subtypes (21). Each cleft subtype was present in each ancestry group, as defined by principal components (PCs) of genetic markers (Table S1). Subjects where the specific subtype of cleft was not known were excluded from this study. Related, affected individuals were retained in this study and a genetic relatedness matrix (GRM) was used to adjust for relationships within and across families (see below).

Samples were genotyped for approximately 580,000 single nucleotide polymorphic (SNP) markers from the Illumina HumanCore+Exome array, of which approximately 539,000 SNPs passed quality control filters recommended by the Center for Inherited Disease Research (CIDR) and the Genetics Coordinating Center (GCC) at the University of Washington (2). These data was then phased with SHAPEIT2 (22) and imputed with IMPUTE2 (23) to the 1000 Genomes Project Phase 3 release (September 2014) reference panel. The most-likely imputed genotypes were selected for statistical analysis if the highest probability (r^2^) > 0.9. SNP markers showing deviation from Hardy-Weinberg equilibrium in European controls, a minor allele frequency or MAF < 5%, or imputation INFO scores < 0.5 were filtered out of all subsequent analyses. A GRM was calculated from a set of LD-pruned genotyped SNPs as defined by GCTA using the package SNPRelate (24).

### Statistical analyses

#### Subtype-specific genome-wide association study (GWAS)

Single subtype genome-wide tests were done by comparing cases from each subtype to a group of unrelated controls to test for genetic variants associated with each cleft subtype. The association between every imputed genetic variant and laterality type was tested using the generalized linear mixed model (GMMAT) (25) as implemented in the GENESIS software package (26). Sex and the estimated GRM were adjusted for under the null model to account for both population substructure and relatedness. SNPs with association p-values less than 5 × 10^−8^ were considered genome-wide significant and those with p-values less than 1 × 10^−5^ were considered ‘suggestive’ and were used for downstream enrichment and comparison analyses. The unadjusted odds ratio (OR) for each SNP was estimated using the minor allele frequency in bilateral cleft cases compared to unilateral cleft cases (27, 28). Regional association plots were made with LocusZoom, where the LD blocks and recombination rates were estimated from European populations (29).

#### Modifier GWAS

We identified genetic modifiers using case-case group comparisons, directly comparing allele frequencies at each SNP between unilateral and bilateral cleft cases. Thus, this approach has high power to identify genetic risk factors that differ between two subtypes, but no power to find factors important in both groups (i.e., SNPs detected in previous GWAS of CL, CLP, or the combined CL/P group) (24). Therefore, this test has the potential to identify new loci for which there is an effect in only one subtype or where the effects are different between two groups; such loci may be masked in an overall scan when the two groups are combined. We performed modifier analyses for severity separately in the CL and CLP subtypes (UCL vs. BCL and UCLP vs. BCLP) and combined as CL/P (UCL/P vs. BCL/P). Similar to the modifier analysis above, these tests were done using GMMAT (25) as implemented in GENESIS (26), adjusting for sex and the GRM to account for both population substructure and relatedness. The OR for each SNP was estimated using the minor allele frequency in cases compared to controls (27, 28). Regional association plots were made with LocusZoom(29).

#### Comparisons between CL and CLP analyses

The estimated ORs for suggestive SNPs (i.e. those with p < 1 × 10^−5^) in the subtype-specific analyses were compared both within a single severity subtype across cleft type (i.e. BCL vs. BCLP), and across severity types within a single cleft type (e.g. UCL vs. BCL). To compare whether the SNPs associated with individual subtypes were novel compared to what has already been reported in previous GWAS of CL, CLP, or CL/P, the SNPs in these analyses within 50kb of previously associated risk SNPs (2, 5, 6, 20) were also identified. A similar approach was done for the modifier analysis, and the suggestive loci, from either the CL or CLP modifier analyses were compared to see if they either had overlapping 95% confidence intervals (CIs) or gave estimated effects in the same direction.

### Replication cohort

To replicate the statistically significant results from our modifier analysis, data from the GENEVA consortium was used, which was described previously (2, 4, 20). Briefly, this cohort recruited case-parent trios, where the affected individual had an oral cleft. The samples were genotyped for approximately 589K SNPs using the Illumina Human610-Quadv.1_B BeadChip, phased using SHAPEIT, and imputed to the 1000 Genomes Project Phase I (June 2011) reference panel using IMPUTE2. Imputed genotype probabilities were converted to most-likely genotype calls with GTOOL (http://www.well.ox.ac.uk/~cfreeman/software/gwas/gtool.html). This dataset was subsequently filtered to only include common SNPs with a MAF > 5%. A subset of individuals was included in both the POFC study and the GENEVA consortium, and these were removed from the replication analysis so that the two groups would be independent. Only the cases from this GENEVA cohort were selected, and they were classified as BCL (N = 28), UCL (N = 326), BCLP (N = 301), UCLP (N = 678). PCs of ancestry were calculated using PLINK (v1.9) (30). Modifier analyses (comparing BCL vs. UCL and BCLP vs. UCLP) were conducted using logistic regression models in PLINK (v1.9), with sex and the first 4 PCs as quantitative covariates. Because of the small sample sizes in the replication cohort and the differences in genotyping arrays and imputation panels, only regions that were significant in the original modifier analysis were tested in this replication strategy. P-values less than a Bonferroni correction for the number of SNPs in the region (0.05/the number of SNPs tested) were considered to be evidence of significant replication.

### Association with normal facial variation

The genome-wide significant modifier locus was further examined in relation to normal facial variation by reviewing the association results of SNPs in this locus in a GWAS meta-analysis of facial shape in two large cohorts (n= 8,246) from the US (MetaUS) and UK (MetaUK) (31). To analyze normal facial variation, a data-driven global-to-local facial segmentation approach was performed. Multivariate GWAS was then performed in each of the resulting 63 hierarchically arranged facial segments. More information on the analysis pipeline and the cohorts can be found in the initial study (31).

### Epigenomic context of results

Topologically-associated domains (TADs) were defined for significantly associated loci using the H1-ESC cell line in 3D Genome Browser (32). Functional enrichment was tested by first annotating all of the SNPs to the craniofacial functional regions defined by Wilderman *et. al*. (33) for human embryos at CS13, CS14, CS15, CS17, and CS20 (4.5-8 weeks post conception). Enrichment tests were done using a chi-square test with the top SNPs (p < 1 × 10^−3^) for both modifier analyses and each subtype analysis, so that group sizes would be sufficient for testing, and estimated ORs and their 95% CIs were calculated.

## Results

We performed a subtype-specific genome-wide analysis for BCL, UCL, BCLP, and UCLP cases by comparing cases of each subtype to unaffected controls. This approach can detect variants associated with increased risk for an OFC in general, but also has the potential to identify variants that increase the risk for one or more subtypes of OFC. A single SNP in chromosome 3q28 achieved genome-wide significance in the analysis of BCL (rs72439195; p = 3.69× 10^−8^), and 90 regions yielded suggestive evidence, most of which have not been previously implicated in OFC formation. However, some of these regions, like 14q32.33 (lead SNP: rs61996057; p = 8.07× 10^−8^; Figure S1A, Figure S2A, Figure S3, Table S2), have been implicated in syndromes with facial dysmorphisms (34-36). In the analysis of UCL, two loci reached genome-wide significance (8q24 and 1q32), both of which are recognized genetic risk loci for CL/P (Figure S1B, Figure S2B, Table S3) (2, 4, 7-10). Among the 21 suggestive loci, 17 have not been previously associated with OFCs which may reflect a lack of GWAS focused specifically on CL. Some of these loci, such as 2q13 (lead SNP: rs6542368; p = 1.06× 10^−7^; Figure S4), are plausible candidates for craniofacial dysmorphism (37). Both BCLP and UCLP have multiple recognized genes/regions, including 8q24 and 17p13, reach genome-wide significance (Figure S1C-D, Figure S2C-D, Table S4, Table S5), and 35 and 41 loci reach suggestive significance, respectively, in this analysis (2, 4, 5, 8, 10, 17).

Because of the apparent differences in suggestive and significant loci in the subtype-specific GWASs, we wanted to characterize similarity or dissimilarity of the overall genetic architectures of UCL, UCLP, BCL, and BCLP. Therefore, we performed pairwise analyses comparing the odds ratios and 95% CIs for SNPs identified as suggestive in the GWAS for each subtype being compared. In the comparison of BCL and UCL SNPs, we found a striking difference in estimated ORs in which 44.03% of 741 SNPs did not have overlapping CIs. A majority of these SNPs originating from the BCL analysis had an OR near 1 the UCL analysis (Figure 1), indicating substantial differences in the genetic architecture of BCL, the more severe group. This was also seen, although to lesser degree when the BCL subtype was compared to BCLP, where the 95% CIs for the estimated ORs did not overlap for the 34.1% of 1181 suggestive SNPs (Figure S5). In contrast, BCLP and UCLP were quite similar, with 94.7% of their 1093 SNPs showing overlapping OR confidence intervals (Figure 1). We also found SNPs with different effects in the subtype-specific analyses were less likely to have been previously reported in analyses of the combined group CL/P (Figure 1). For example, in the BCL-UCL comparison, 26.8% of SNPs with overlapping estimated effect sizes were recognized CL/P risk SNPs, indicating these SNPs may predispose to OFC risk but have no effect on specific subtypes. However, only 1.8% of SNPs differing in their effect sizes were previously reported, significantly less than expected by chance alone (p = 2.41× 10^−20^). This pattern held for all comparison groups (Table S6). We reasoned that SNPs predisposing to any type of bilateral cleft could be identified by first selecting SNPs that had non-overlapping CIs between BCL and UCL that also had overlapping CIs between BCL and BCLP. However, only 4 SNPs met these criteria and all of them also showed nominal significance in UCLP and had overlapping CIs. We employed the same strategy to identify SNPs predisposing to any type of unilateral cleft, but were similarly unsuccessful, supporting the notion that subtype-specific risk factors are not shared between CL and CLP in this sample.

**Figure 1:**
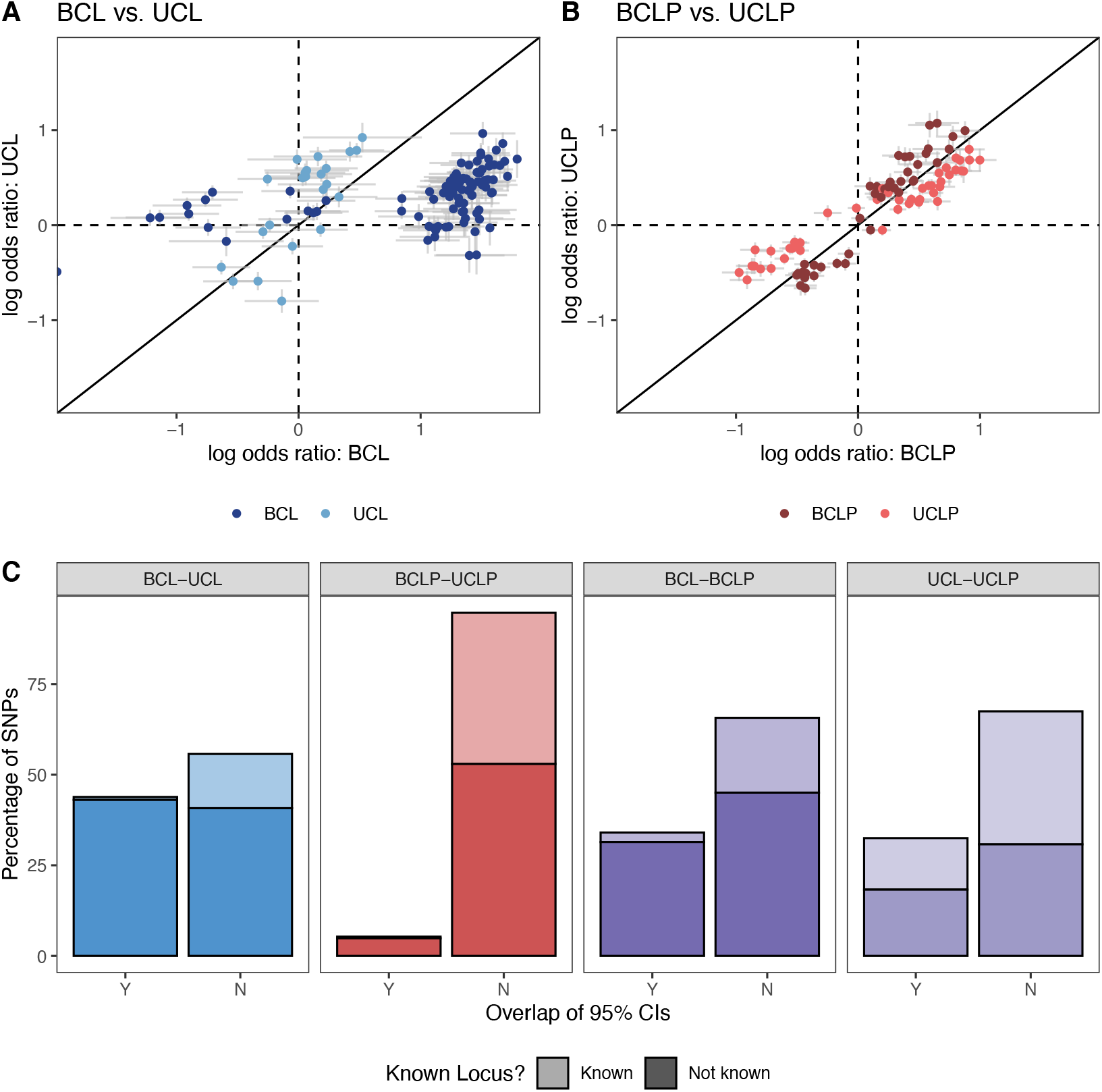
The log odds ratio for SNPs that were suggestive (p < 1 × 10^−5^) or significant (p < 5 × 10^−8^) in the subtype-specific case-control analyses in were compared between BCL and UCL (A), BCLP and UCLP (B), and were classified by whether the confidence interval for the odds ratio overlapped and whether the variant was known (C).

To disentangle the effects of SNPs on specific subtypes from more general effects on OFC risk, we performed a genome-wide bilateral vs. unilateral modifier analysis in CL and CLP cases. Because this is a case-to-case group comparison, this analysis would not be able to detect variants generally important for both CL or CLP risk, but would detect variants important for the formation of one severity subtype compared to the other. In the modifier analysis of CL, one locus on chromosome 20p11 reached genome-wide significance (lead SNP: rs143865354; p = 7.53×10^−9^) and 47 other SNPS yielded suggestive significance (Figure 2A; Table S7; Figure S6A). In the modifier analysis for CLP, no loci reached genome-wide significance, but 19 loci yielded suggestive significance (Figure 2B; Table S8; Figure S6B). Interestingly, when CL and CLP were combined (as is typical in genetic analyses of OFCs), no loci reached genome-wide significance, and only 3 loci gave suggestive significance (Figure S7; Table S9), raising the possibility that these modifiers may not be shared between CL and CLP.

**Figure 2:**
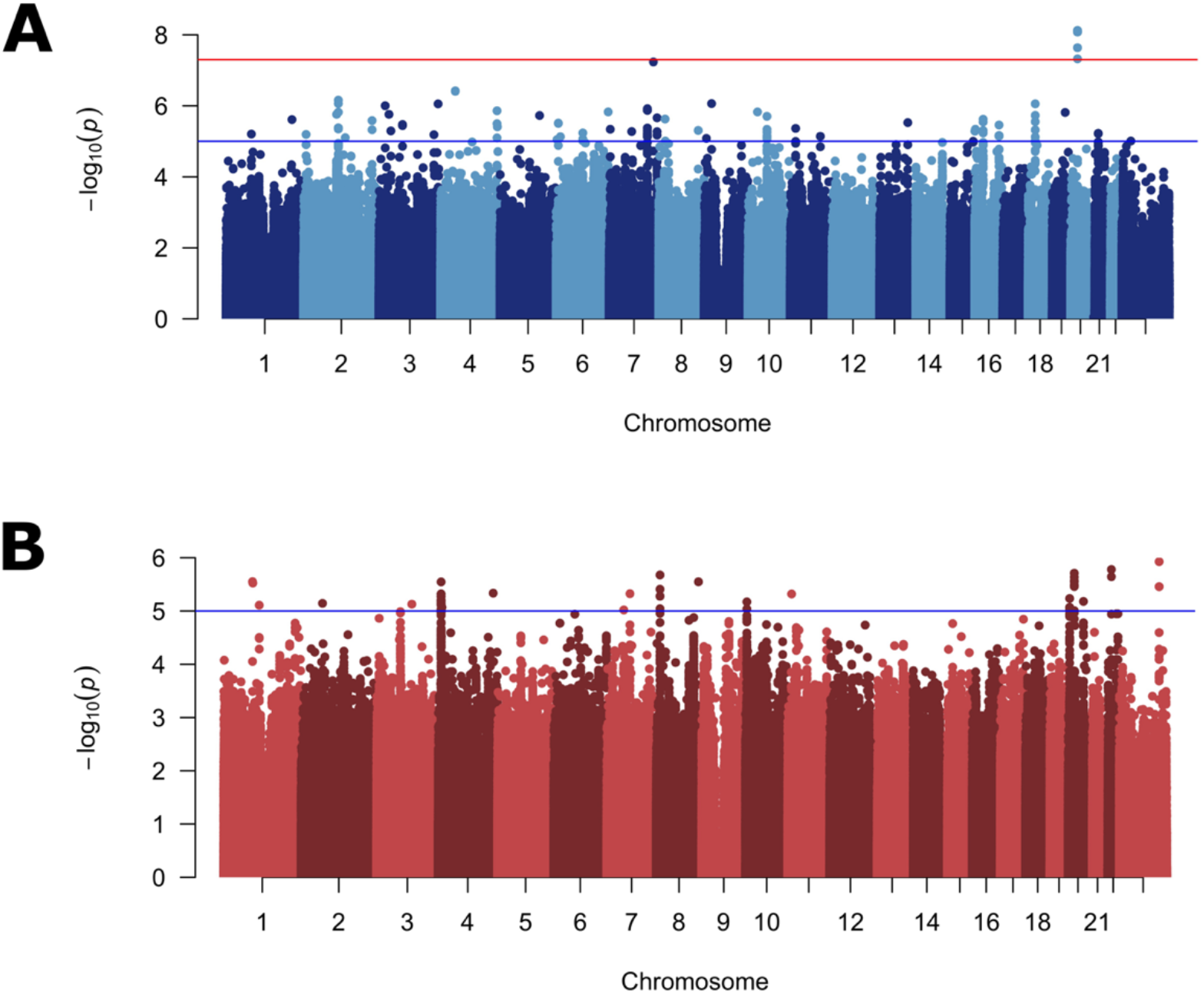
Manhattan plots of –log_10_(p-values) from the bilateral vs. unilateral modifier analysis in participants with cleft lip (A), and cleft lip and palate (B). Lines indicate suggestive (blue) and genome-wide (red) thresholds for statistical significance. The genomic inflation factors were 0.96 and 1.01, respectively.

The associated SNPs on 20p11 lie within *LINC01432*, and are within the same topologically associated domain as *PAX1* (Figure 3A; Figure S8). This locus was not significant (p > 0.05) in the modifier analysis of CLP (Figure 3B). Additionally, when the OR for the lead SNP in this region was compared between CL and CLP cases, the direction of effect was not consistent (with either a 95% CI or a 99% CI; Figure 3C). We replicated the 20p11 region in an independent sample of 28 BCL cases, 329 UCL cases, 306 BCLP cases, and 685 UCLP cases. In this 20p11 region, there were 8 SNPs passing filtering in the CL modifier analysis.

**Figure 3:**
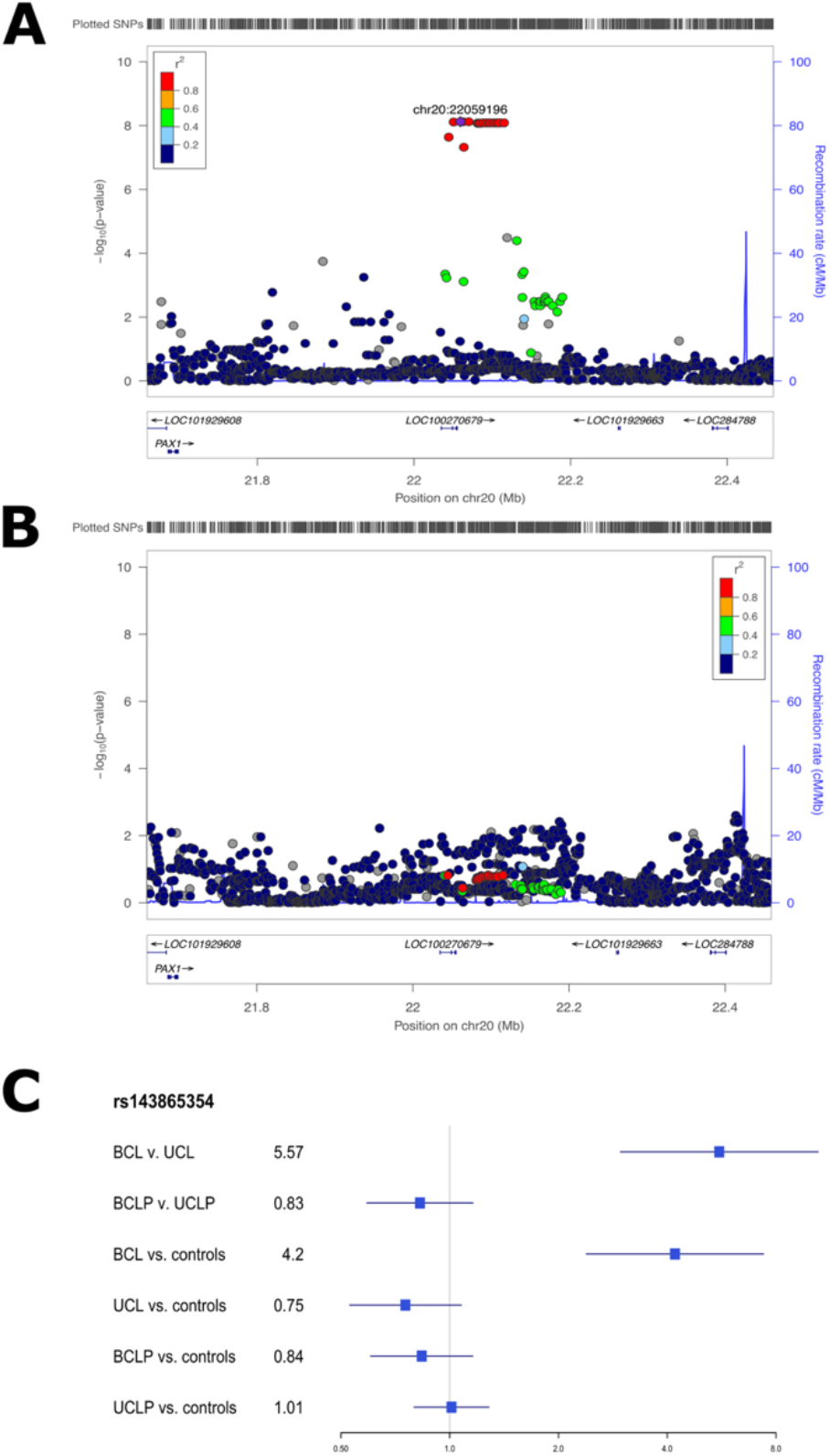
Regional association plots showing –log_10_(p-values) for the novel genome-wide significant peaks at 20p11 in the modifier analysis in cleft lip (A) and cleft lip and palate (B). Plots were generated using LocusZoom (29).The recombination overlay (blue line, right y-axis) indicates the boundaries of the LD block. Points are color coded according to pairwise LD (r^2^) with the index SNP. The odds ratio for this locus in each of the modifier and subtype specific loci were also compared (C).

While none of these SNPs were the same as those in the original analysis, one SNP (rs28970569) was also a significant modifier in the replication cohort (OR = 3.83, 95% CI = 1.64-8.95, p = 0.0018; Table S10). In the CLP modifier analysis, 9 SNPs passed our filters but none of were significant modifiers, consistent with the results for 20p11 in our discovery sample (p > 0.05; Table S11). Additionally, we wanted to determine the extent to which the genetic modifiers in CL were similar to the genetic modifiers in the CLP genome-wide analysis. To test this, we compared SNPs that were suggestive (p < 1 × 10^−5^) in either the CL or CLP modifier analyses. Notably, there was no overlap between the list of suggestive SNPs in CL and the list of SNPs suggestive in CLP. Moreover, the estimated ORs were not positively correlated and all of the suggestive SNPs in the analysis of CL had no effect in CLP and *vice versa* (Figure 4), and a majority of the SNPs in each analysis were not near regions previously associated with CL/P (Table S12). Cumulatively, these results suggest the 20p11 modifier for bilateral vs. unilateral OFCs is specific to CL.

**Figure 4:**
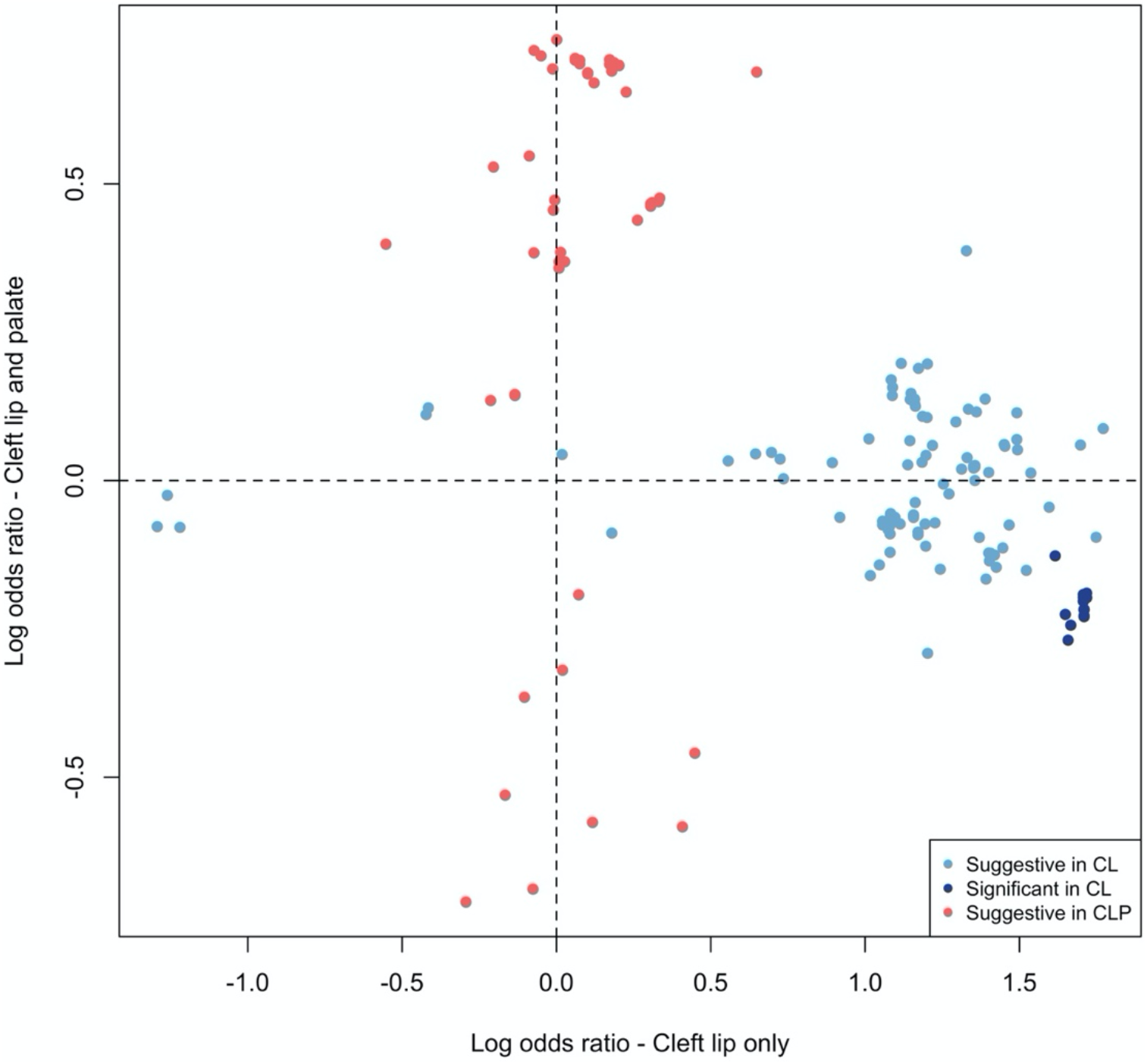
The log odds ratios for 188 SNPs that were suggestive (p < 1 × 10^−5^) or significant (p < 5 × 10^−8^) in the modifier analysis in CL or CLP were compared. No SNPs were genome-wide significant in CLP. No SNPs were significant or suggestive in both CL and CLP.

Although the 20p11 locus had not previously been associated with risk to OFCs, it has been associated with variation in normal facial structures. Therefore, we next investigated whether the BCL modifier SNPs were also associated with normal facial variation as that could give insights into how these SNPs might influence cleft severity. We found that rs6036034, a SNP in the 20p11 region in LD with rs143865354 (R^2^ = 0.522; p= 4.75× 10^−8^ in BCL vs. UCL), was associated with normal variation in nose morphology (p = 2.63× 10^−11^), specifically projection of the nasal tip and columella and breadth of the nasal alae (Figure 5). These are the same structures disrupted by cleft lip and are derived from the lateral nasal processes where *PAX1* is expressed (38). Moreover, rs143865354 shows modest evidence of being an eQTL for *PAX1* in skin (p=2.9× 10^−5^) in GTEx.

**Figure 5:**
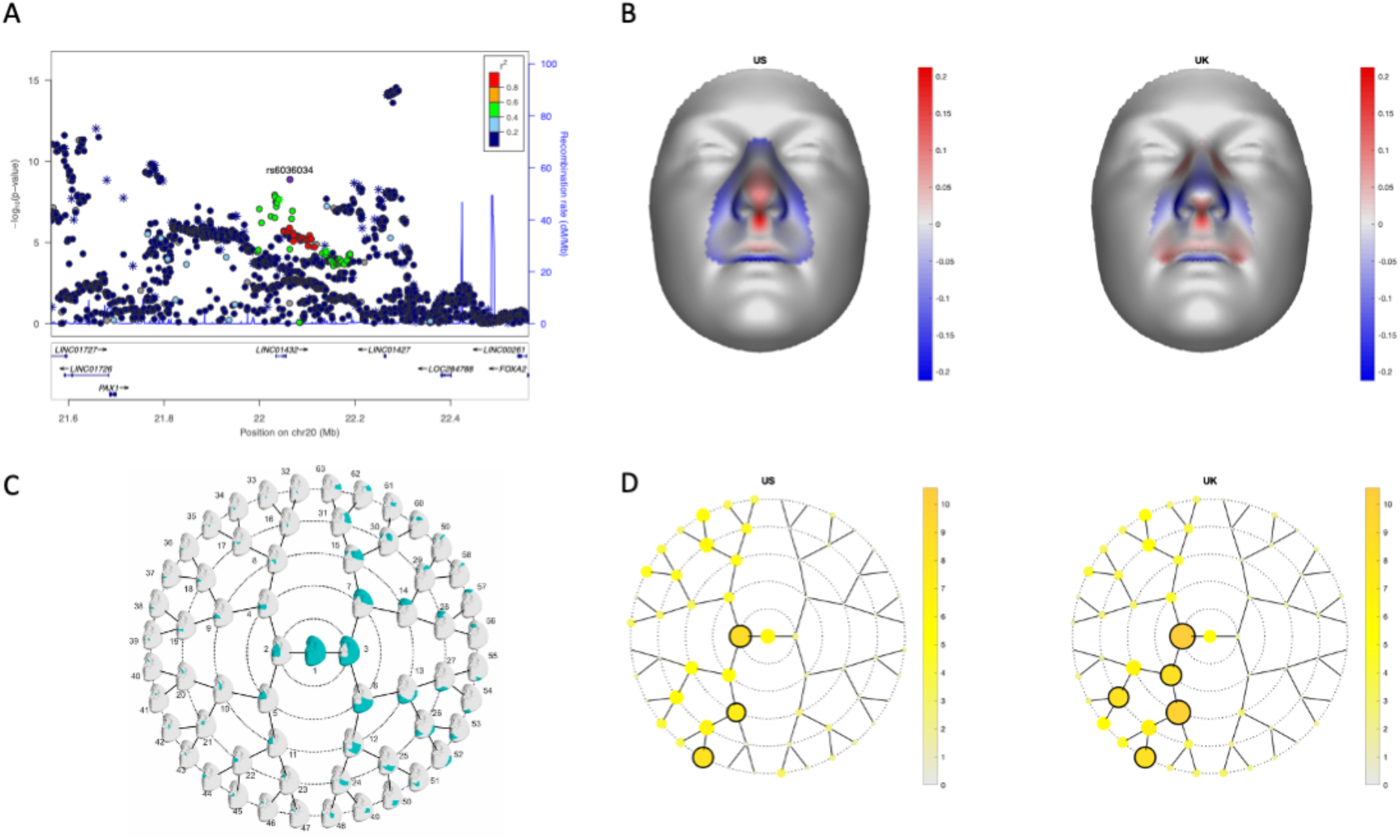
LocusZoom plots for the association of normal facial variation and rs6036034 (A). Points are color coded based on linkage disequilibrium (R^2^) in Europeans. The asterisks represent genotyped SNPs, the circles represent imputed SNPs. The normal displacement (displacement in the direction locally normal to the facial surface) in each quasi-landmark of the facial segment reaching the lowest p-value in MetaUS and MetaUK going from the minor to the major allele SNP variant (B). Blue, inward depression; red, outward protrusion. Global-to-local facial segmentation plot that shows the 63 facial segments represented in teal obtained using hierarchical spectral clustering (C). The -log10(p-value) of the meta-analysis p-values per facial segment in MetaUS and MetaUK. Black-encircled facial segments have reached a genome-wide p-value (p = 5.00×10^−8^) (D).

We were also interested in testing whether differences in genetic architecture in BCL, UCL, BCLP, and UCLP at the SNP level were also reflected in functional elements involved in facial development. Therefore, we tested whether SNPs associated with each subtype were enriched in similar functional regions defined by epigenetic marks in human embryonic craniofacial tissues (33). For some elements, the apparent enrichment or depletion was consistent across subtypes. For example, BCL, UCL, BCLP, and UCLP SNPs were similarly depleted in heterochromatin regions, and most were enriched in regions of strong transcription. However, there were some regions showing opposite enrichments in the different subtypes. For example, zinc finger repeat regions were enriched in both BCLP and UCLP, but were depleted in BCL (Figure 6). Interestingly, the severity modifiers for both CL and CLP were depleted in regions of weak transcription, and enriched in regions of low activity. Some of the suggestive modifier loci for CLP were enriched in bivalent transcription start sites but none of the putative modifiers for risk to CL were enriched in functional domain. These enrichment/depletions were consistent throughout craniofacial development (4.5-8 weeks post conception; Figure S9; Table S13). These observations, while not definitive, lend some support the idea that although at the SNP level, the genetic underpinnings for cleft subtypes are distinct, this may not extend entirely to gross differences in functional element enrichments. Deciphering the true underlying mechanism(s) resulting in bilateral and unilateral CL and CLP will require a locus-by-locus investigation.

**Figure 6:**
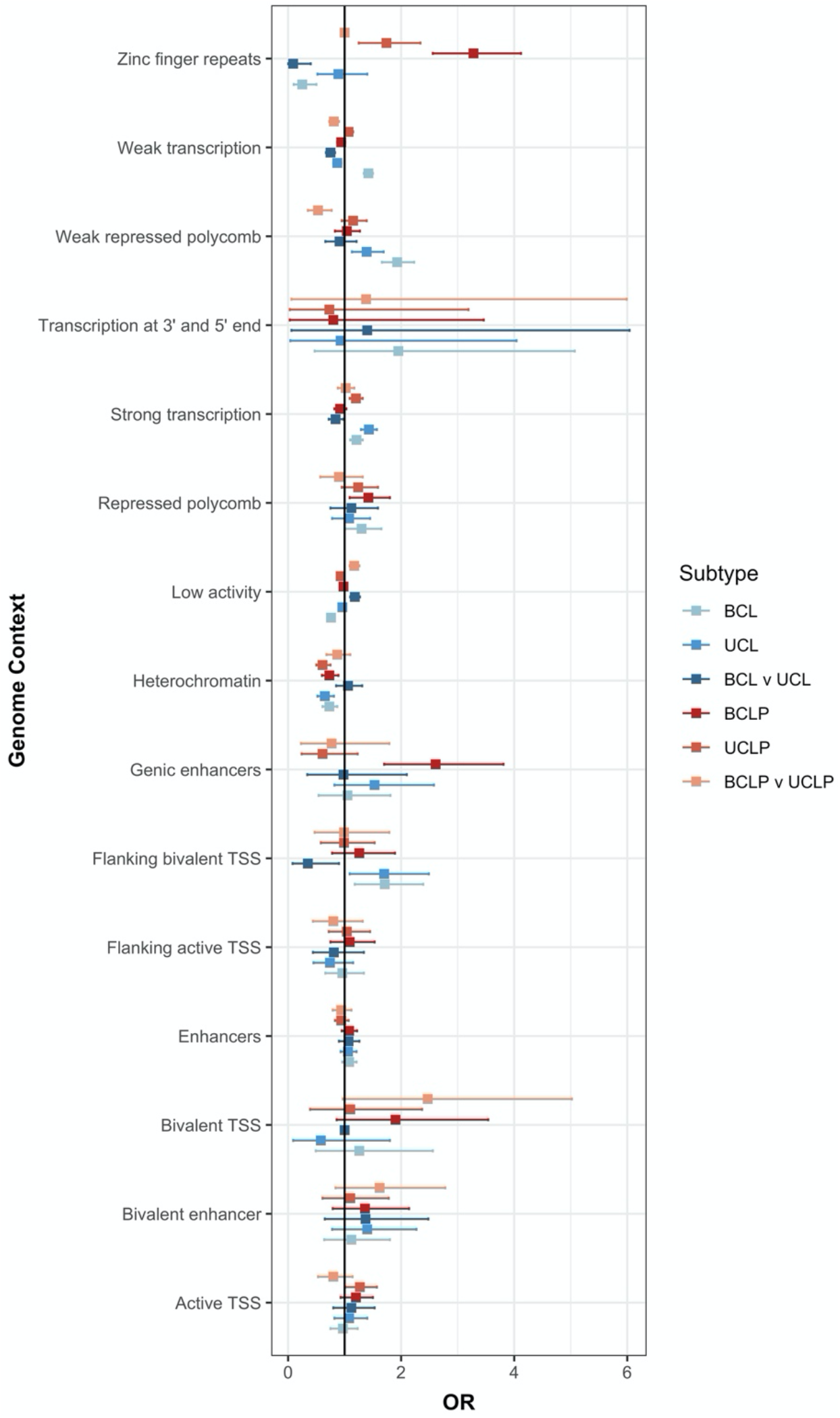
Enrichment of the top SNPs associated in either the CL modifier analysis, CLP modifier analysis, and each subtype analysis (p < 1 × 10^−3^) were tested in each functional region defined during craniofacial development (CS15).

## Discussion

While there have been many studies identifying genetic variants that influence overall risk to CL/P and CP, the genetic underpinnings of specific phenotypic subtypes of cleft lip is less studied. This report furthers our understanding of genetic variants associated with specific subtypes of OFC: BCL, UCL, BCLP, and UCLP. We used a modifier analysis which provides more power to find genetic loci differing between two groups, and found one locus on 20p11 that replicated in an independent cohort as significantly associated with the formation of a BCL over a UCL. The associated SNPs were located in several long, noncoding RNAs and within the same TAD (300kb downstream) as the *PAX1* gene. While *PAX1* has not been associated with OFC like its paralog *PAX9* (39), they both are transcription factors with similar DNA-binding domains regulating chondrocyte differentiation and the formation of invertebrate discs, and knock-out mouse models show skeletal abnormalities (40-42). There is also evidence that *PAX1* is upregulated by *SHH*, and in turn, upregulates *SOX5* and *BMP4* (41-43). There is only a limited literature describing P*AX1* expression in the developing face (38) and it has not been previously associated with risk to nonsyndromic OFCs, but *PAX1* is in a pathway with other genes known to be associated with nonsyndromic OFCs (44-49). Additionally, recent studies have shown mutations in *PAX1* cause otofaciocervical syndrome (OTFCS, MIM#615560) which presents with facial dysmorphisms (50, 51), and studies of normal facial variation have found this locus has also been associated with nasal width (the distance between left and right cartilaginous nasal ala) in people of European descent (52) and in people of Latin American descent (53). The link between SNPs at the *PAX1* locus and normal facial shape was further substantiated in our analysis, with effects observed in the nasal tip, columella and alae. These anatomical structures are derived from the lateral and medial nasal processes in the embryo, which form the primary palate. Thus, it is biologically plausible that *PAX1* could affect the development of specific types of craniofacial abnormalities, however, more work is be needed to investigate the underlying mechanisms.

While 20p11 was the only genome-wide significant modifier found in this study, this may partly be due to limited sample size in some of the OFC subtypes. It is important to note that when a modifier analysis was conducted on all combined CL and CLP cases, fewer loci reached even suggestive significance, suggesting CL and CLP may have distinct modifiers. Consistent with this, the suggestive modifiers for risk in CL and CLP showed no overlap in estimated effect on risk. This suggests that the lack of overlap is not entirely due to a difference in sample size, but that instead that there is a biological difference in the genetics of laterality in CL compared to CLP.

This is the first study to test for severity modifiers at a genome-wide level, but we previously tested for modifiers in 13 recognized GWAS regions known to be associated with OFCs (54) and found SNPs in *IRF6* were associated with the formation of a unilateral CL/P compared to bilateral CL/P (17). In our study, no SNPs in *IRF6* reached suggestive significance. Our study was larger than the previous study (2339 cases vs. 1001 cases), therefore, this difference may reflect effects of modifiers for cleft subtypes in regions of genome not recognized by previous GWAS of OFCs. This is not surprising given OFC subtypes are typically combined for GWAS, which maximizes statistical power to detect loci associated with overall risk, but would mask loci with different effects in subtypes.

We also conducted analyses comparing each subtype to unrelated controls. This analysis should find loci associated with either overall risk or one particular cleft subtype, but would have less statistical power to detect loci that differ between two subtypes. Most loci achieving genome-wide significance in these analyses were those already recognized to be associated with risk to OFCs (2, 5, 6, 20). There were, however, some loci yielding suggestive evidence of association for several of the subtype-specific analyses not previously reported, but could be in the causal pathway for syndromes with facial dysmorphisms. For example, SNPs in 14q32.33 gave suggestive evidence of association for BCL, with a distinct effect only seen in BCL, and 2q13 yielded suggestive evidence of association for UCL. Microdeletions in both of these regions have been associated with syndromes that include facial dysmorphisms (34-37). The 14q32.33 also contains *JAG2* which is part of the Notch signaling pathway and is important for craniofacial development (55-57).

Overall, our analyses demonstrated that BCL was most distinct from the other three subtypes analyzed and that these modifiers were not shared between CL and CLP. We found that the associated SNPs in all four OFC subtypes were enriched in regions associated with transcription and depleted in heterochromatin regions. This was expected because nonsyndromic OFCs form from the disruption of one of the processes involved in facial development and thus variants associated with any subtype OFC should be enriched in regions active during facial development. It is also consistent with the study defining the functional regions, which showed enrichment in active states for SNPs involved in overall OFC risk (33). Importantly, there were some differences in functional enrichment by subtype. For example, SNPs associated with BCLP and UCLP were enriched in zinc finger repeat regions, however, SNPs showing some evidence of association with BCL were depleted in this same region. This further emphasizes the possibility for a distinct genetic architecture associated with risk to BCL. Additionally, the modifiers for both CL and CLP were depleted in regions associated with active transcription and strongly enriched in regions of low activity. This result is somewhat surprising given it is the opposite of what would be expected for an analysis involving craniofacial development. However, the biological mechanism by which modifiers could affect a phenotype is not known. Therefore, this highlights the need for more studies that test how modifiers mechanistically act.

The findings from this study should also consider its limitations. Many of the subtypes of clefting, particularly BCL, had small sample sizes. Limits of small sample sizes make it likely other subtype-specific genetic loci and modifiers may exist and we are unable to detect them in this statistical analysis. Additionally, we were unable to test for heterogeneity across ancestry groups while testing for subtype-specific genetic risk loci and severity modifiers. This cohort is multiethnic, including people of European, Asian, and Latin American ancestry, and previous studies have shown ancestry-specific association with risk to OFC (2). Studies with larger sample sizes for these clefting subtypes could lead to the discovery of more associated genetic loci and test for differences in associated loci between different ancestry populations.

In summary, we conducted the first genome-wide scan for severity modifiers in a case-case and case-control design focused on nonsyndromic CL and CLP and found a significant modifier in 20p11 downstream from *PAX1* associated with increased risk for BCL over UCL. We also showed these modifiers for CL and CLP were distinct, with the modifiers of one cleft sub-type have little to no genetic effect in the other subtypes. Furthermore, in the subtype-specific GWASs, we found several suggestive loci that had not been previously identified in previous GWASs that combined cleft subtypes. We also found loci associated with BCL were the most distinct from those associated with other cleft subtypes, suggesting the etiology of this rarest subtype of cleft to be unique. Overall, this study expands our understanding of the genetic underpinnings of the genetic and phenotypic heterogeneity of OFCs and suggests new areas of research on cleft lip subtypes.

## Data Availability

All of the genetic data used in this study was previously generated and is available on dbGAP

## Acknowledgments

The authors thank the dedicated field staff, collaborators, and participating families for their important contributions to this study. This work was supported by grants from the National Institutes of Health (NIH) including: R00-DE025060 [EJL], X01-HG007485 [MLM, EF], R01-DE016148 [MLM, SMW], U01-DE024425 [MLM], R37-DE008559 [JCM, MLM], R01-DE009886 [MLM], R21-DE016930 [MLM], R01-DE012472 [MLM], R01-DE014581 [TB], U01-DE018993 [TB]. National Institute of Dental and Craniofacial Research (U01-DE020078; R01-DE027023 [SMW]). Funding for genotyping by the National Human Genome Research Institute (X01-HG007821) and funding for initial genomic data cleaning by the University of Washington provided by contract HHSN268201200008I from the National Institute for Dental and Craniofacial Research awarded to the Center for Inherited Disease Research.

